# The epidemiology of autoimmune liver disease varies with geographic latitude

**DOI:** 10.1101/2020.06.27.20141382

**Authors:** Gwilym J Webb, Ronan P Ryan, Tom P Marshall, Gideon M Hirschfield

## Abstract

**Background & aims:** The epidemiology of autoimmune liver disease (AILD) is challenging to study because of the diseases’ rarity and because of cohort selection bias. Increased incidence further from the Equator is reported for multiple sclerosis, another autoimmune. We assessed the incidence of primary biliary cholangitis (PBC), primary biliary cholangitis (PSC), and autoimmune hepatitis (AIH) in relation to latitude.

**Methods:** Retrospective cohort study using anonymised UK primary care records 2002-01-01 to 2016-05-10. All adults without a baseline diagnosis of autoimmune liver disease were included and followed until first occurrence of an AILD diagnosis, death, or they left the database. Latitude was measured as registered general practice rounded down to whole degrees.

**Results:** The cohort included 8 590 421 records with 53.3 10^7^ years follow-up from 694 practices.

There were 1314 incident cases of PBC, 396 of PSC, and 1034 of AIH. Crude incidences (95% confidence interval) was: PBC 2.47 (2.34-2.60), PSC 0.74 (0.67-0.82), and AIH 1.94 (1.83-2.06)/100 000/year. PBC incidence correlated with female sex, smoking, and deprivation; PSC incidence correlated with male sex and not smoking; AIH incidence correlated with female sex and deprivation.

More northerly latitude was strongly associated with incidence of PBC: 2.16 (1.79-2.60) to 4.86 (3.93-6.00) from 50-57°N (p=0.002) and AIH 2.00 (1.65-2.43) to 3.28 (2.53-4.24)(p=0.003), but not PSC 0.82 (0.60-1.11) to 1.02 (0.64-1.61)(p=0.473).

Incidence after adjustment for age, sex, smoking, and deprivation status showed similar positive correlations for PBC and AIH with latitude, but not PSC. Incident AIH cases were younger at greater latitude.

**Conclusions:** We describe a novel association between increased latitude and the incidence of PBC and AIH that requires both confirmation and explanation.

## Introduction

The three major autoimmune liver diseases (AILDs), primary biliary cholangitis (PBC), primary sclerosing cholangitis (PSC), and autoimmune hepatitis (AIH), represent significant causes of liver morbidity and mortality whose aetiopathogenesis is incompletely understood.[1–3] Although individually uncommon, together they account for a significant proportion of elective liver transplantation,[4] and incidence and prevalence appear to be increasing.[5–8]

To date, genetic association studies have only been able to explain a minority of the risk for the three AILDs, suggesting a significant role for environmental factors.[9] A number of environmental factors have been identified as associated with risk of AILD including urinary tract infections, nail polish use, hair dye use, smoking, and deprivation in PBC [10–14]; smoking as a negative risk factor in PSC [14–16]; and various drug exposures in AIH.[3, 17, 18] However, even in aggregate, these factors appear insufficient to explain variations in individual disease risk.[9]

Latitude has been proposed as a risk factor for AILDs because of its link to vitamin D status and in turn the link between vitamin D status and risk of autoimmunity.[19–23] T_H_1 T cell activation and B cell activation are implicated in each of the AILDs and activation is negatively regulated by physiological concentrations of vitamin D.[1–3, 22] The association between the neurological autoimmune disease multiple sclerosis (MS) and latitude is well established.[24, 25] For type 1 diabetes, childhood hypovitaminosis D has been associated with increased disease risk.[26] By analogy, similar associations with latitude exposure have been demonstrated in other autoimmune conditions and latitude has been suggested as a target for investigation among the AILDs, but to date such an assessment has not been performed.[21, 27]

In this study we investigate the relationship between latitude and incidence of AILD in a large UK primary care database with established generalisability to the national population.[28, 29] We also describe disease prevalence and assess disease associations with age, sex, deprivation, smoking, and ethnicity.

## Methods

### Study design and population

A retrospective cohort study was undertaken using the anonymised primary care health records contained in the The Health Improvement Network (THIN) database between 2002-01-01 and 2016-05-10.[30] THIN is a United Kingdom based primary care database containing routinely collected electronic patient records. At each consultation, general practitioners record details of the medical encounter, including diagnosis. Demographic details such as age, sex and linked deprivation scores also form part of the electronic record.

### Inclusion criteria

Patients of all ages registered with a practice contributing to THIN during the study period.

### Exclusion criteria

Patients with a previous diagnosis of AILD at baseline were excluded. Patients with potential overlap of autoimmune liver diseases were excluded (see below).

### Follow up

Patients were followed until death, leaving a contributing practice, the date that a practice stopped contributing to THIN or the end of the study period, whichever was latest.

### Exposure

Latitude was measured by geocoding latitude from the postal code of the general practice concerned. This was performed by THIN to avoid identification of individual practices – the locations of individual practices were not available to the authors. Latitude was rounded down to the nearest integer. Because of the small population resident in the 58° latitude band, those resident at 58° were combined with those resident at 57° for analysis. Townsend quintiles were calculated from postcode of place of residence by THIN. Covariable data on age, sex, smoking status, and ethnicity was collected from general practice records. The most recently submitted data in each category at the end of follow-up was used in each case.

### Outcomes

Diagnoses of AILDs were defined using diagnostic codes (Table S1). For PBC and PSC, a single diagnostic code specific to the respective condition was identified; for AIH, a combination of three potential codes validated in a previous study was used.[31] For use as positive and negative controls of conditions with and without published associations with latitude, cases of MS and hypertension were examined using standard diagnostic codes (Table S1).

Cases with two or more lifetime diagnoses of different AILDs (i.e. cases of possible autoimmune overlap) were not considered to represent incident disease for any AILD diagnosis. Cases were considered incident if the date of their first recording was more than one year after the individual’s registration at the practice concerned and also one year after the practice had achieved acceptable mortality reporting.[32] This was to prevent pre-existing diagnoses appearing incident in new registrations at a particular general practice.[33]

Data regarding Townsend deprivation quintile were provided by THIN and derived from postal code of patient residence. Latitude bands were provided by THIN at special request and were derived from the practice postal code.

All data used were those most recent at the end of follow-up for a given individual.

### Statistical analyses

All data were analysed using StataMP v15.2 (StataCorp, Texas, USA) using the Birmingham BlueBEAR high performance computer cluster. We present descriptive statistics, univariable analysis of associations between risk factors and incidence, and multivariable analysis with adjustedments for sex, age, smoking status, Townsend deprivation quintile, and latitude. Where data were adjusted for co-variables, direct standardisation was used. Where direct standardisation was used to adjust for multiple co-variables, individuals with missing data in any category were excluded. For the examination of trends over time, latitude, or quintiles of deprivation, least squares linear regression was performed. For assessing for changes in sex ratios, the *x*^2^ test for trend was used.

Maps were produced using QGIS v3.49 (https://www.qgis.org/) using public domain shapefiles from the UK Ordnance Survey OpenData project.

### Ethical approval

THIN has previously received approval for research using its database from the NHS South-East Multi-Centre Research Ethics Committee in 2003. This study received approval from the THIN Scientific Review Committee: reference 16THIN055.

## Results

Overall, a total of 8 590 421 pseudonymised patient records with a total of approximately 53.3 million years follow-up from 694 practices were examined. First entry to follow-up was 2002-01-01 and the last data collection point 2016-05-10. Median follow-up was 5.2 years (IQR 1.9-10.2). 1314 incident cases of PBC, 396 incident cases of PSC, and 1034 incident cases of AIH were identified.

### Incidence and prevalence

Summary details of incident cases are provided in Tables 1 and S2. When assessed by linear regression, there was no significant change in the incidence of any of PBC, PSC, or AIH over the study period (Table S4 and Figure S1).

**Table 1:**
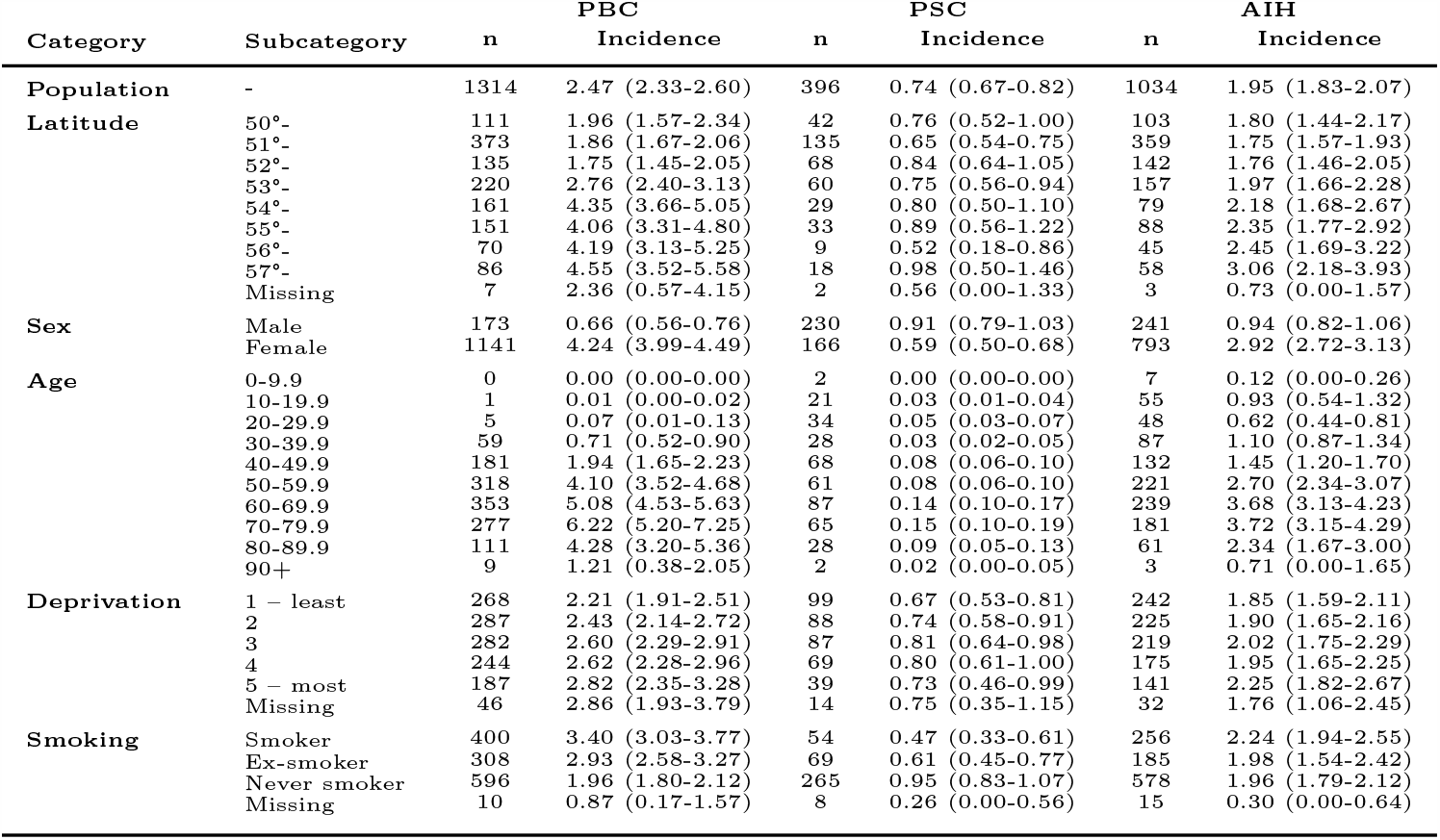
Incidence per 100 000 per year of primary biliary cholangitis (PBC), primary sclerosing cholangitis (PSC), and autoimmune hepatitis (AIH) for the time period 2002-01-01 to 2016-05-10. Figures are adjusted for latitude, sex, age, Townsend deprivation quintile, and smoking status by direct standardisation as appropriate. Figures are per 100 000 population/year with 95% confidence intervals. Deprivation refers to Townsend Deprivation Quintiles. See also Table S2.

In 2015, the last full year of the study, a total of 1299 prevalent cases of PBC, 353 cases of PSC and 1116 cases of AIH were identified. Details of disease prevalence are summarised in Tables 2 and S3. The prevalence of all three diseases increased over time (Figure S2).

**Table 2:**
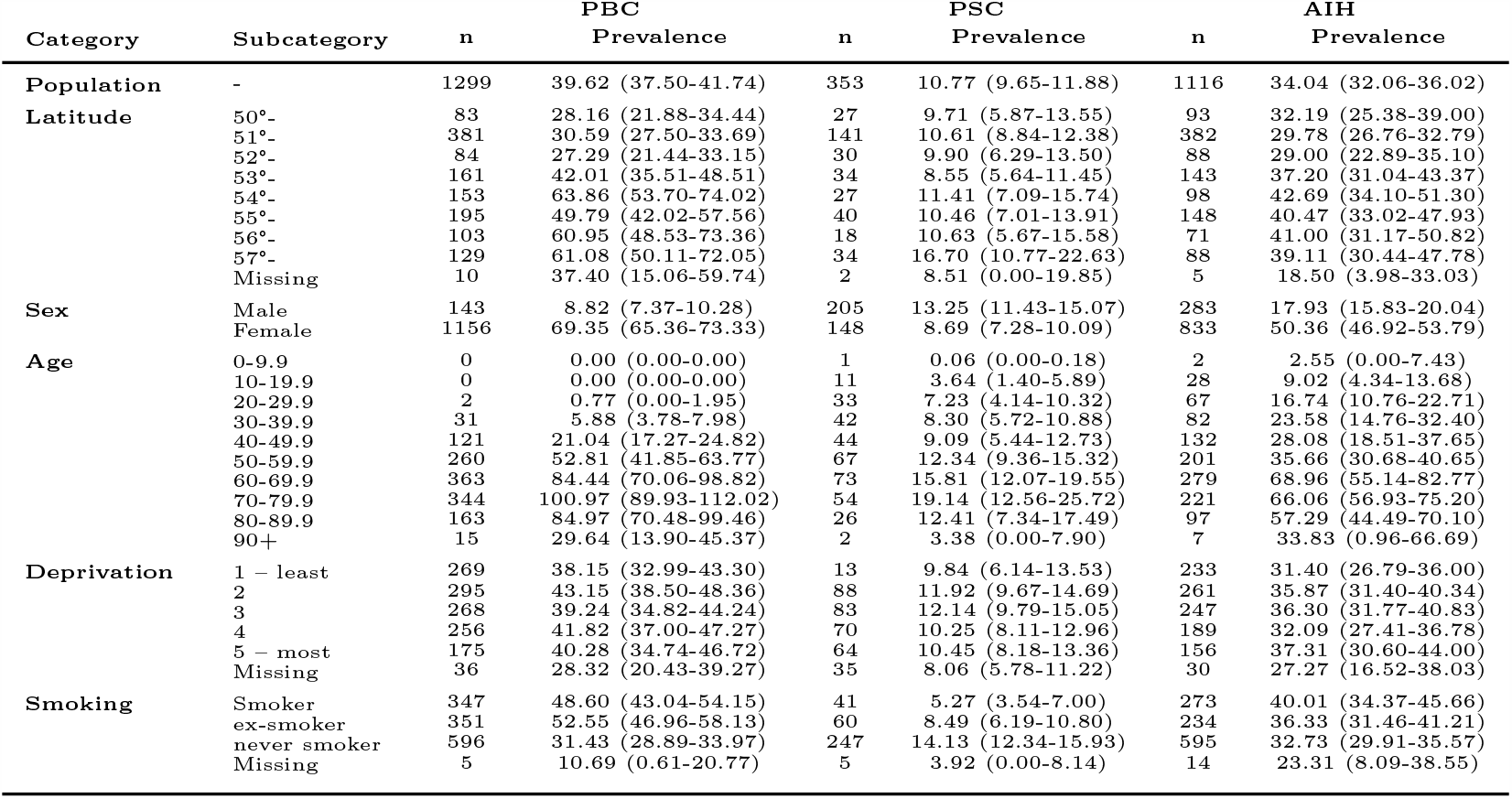
Prevalence of primary biliary cholangitis (PBC), primary sclerosing cholangitis (PSC), and autoimmune hepatitis (AIH) for 2015. Figures are adjusted for one or all of latitude, sex, age, Townsend deprivation quintile, and smoking status by direct standardisation as appropriate. Figures are per 100 000 population with 95% confidence intervals. Deprivation refers to Townsend Deprivation Quintiles. See also Table S3.

### Latitude

To confirm whether a previously reported correlation of incidence with latitude could be demonstrated using this dataset, the incidence of MS was assessed. The overall crude incidence of MS was 8.98 (8.75-9.23)/100 000/year.

When assessed by latitude band after adjustment for sex, age, smoking status, and Townsend quintile, there was a higher incidence of MS in the in the 57° band at 13.67 (11.86-15.48)/100 000/year than in the 50° latitude band at 8.24(7.40-9.07)/100 000/year (Figure S3 and Table S5). There was an increase in MS incidence of 0.66 (0.25-1.07)/100 000/year per degree in latitude increase (r^2^=0.721; p=0.008). By contrast, for hypertension, a disease not reported to have an association with latitude, overall incidence was 946.26 (943.58-948.94)/100 000/year with no significant correlation with latitude at −5.02(−30.27 to 20.23)/100 000/year per degree in latitude r^2^=0.038 and p=644. (Figure S3 and Table S5).

For AILDs, the crude incidence of PBC was markedly greater at more northerly latitudes (Tables 1 and S2, Figure 1, Figure 2, and Figure S4). After adjustment for sex, age, smoking status, and Townsend deprivation quintile there remained a more than doubling in incidence from the 1.96 (1.57-2.34)/100 000/year in the 50° latitude band to 4.55 (3.52-5.58) in the 57° latitude band. When assessed by linear regression, PBC incidence increased by 0.46 (0.27-0.66)/100 000/year per degree in latitude; r^2^=0.850; p=0.001. Similarly, but less markedly, the incidence of AIH was greater at more northerly latitudes at 0.19 (0.11-0.26)/100 000/year/degree; r^2^=0.873; p*<*0.001. PSC incidence showed no significant correlation with latitude: 0.01 (*-*0.02-0.04); r^2^=0.055; p=0.577.

**Figure 1:**
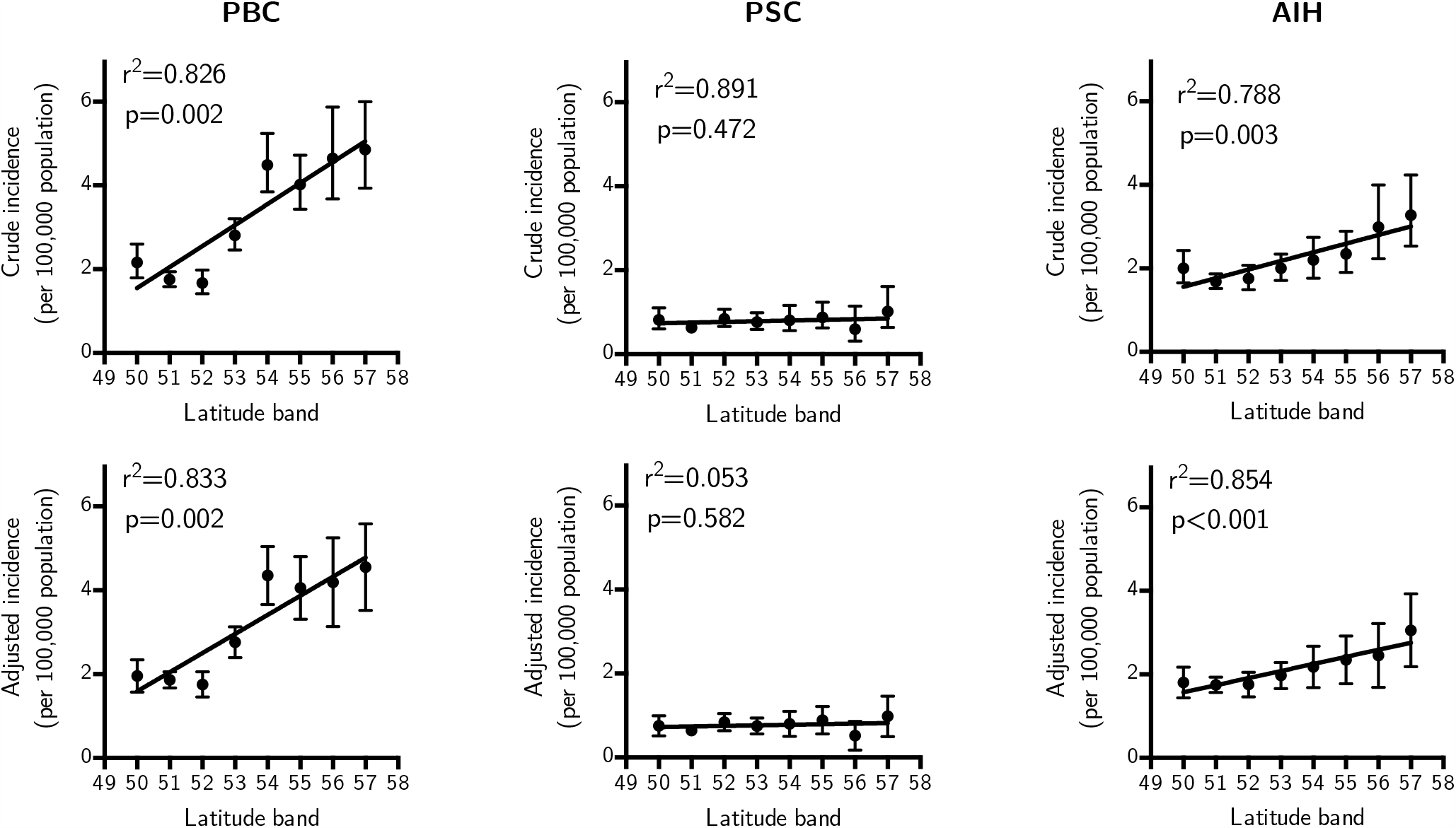
Incidence of autoimmune liver disease by latitude. Plots denote crude incidence (top row) of PBC, PSC, and AIH and adjusted incidence (bottom row) after adjustment for sex, age, smoking status, and Townsend deprivation quintile. For PBC and for AIH, there was a significant increase in incidence at more northerly latitudes both before and after adjustment; for PSC a significant correlation was not present.

**Figure 2:**
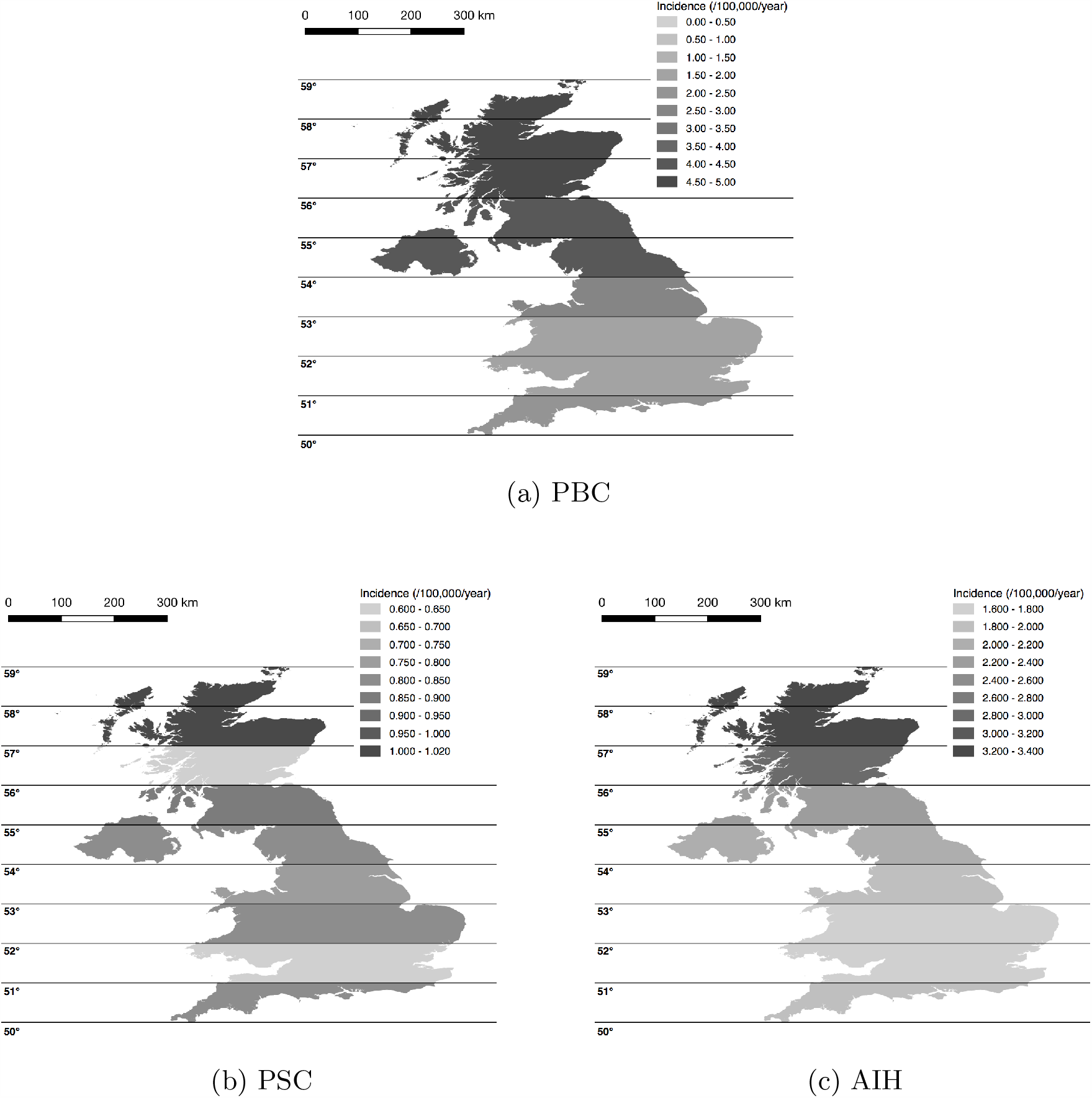
Maps of United Kingdom showing crude incidence of autoimmune liver diseases. Incidences are for the whole study period and given as cases/100 000/year. Density of shading corresponds to incidence as denoted in each panel.

In 2015, after adjustment for age, sex, smoking status and Townsend quintile, there was a significant increase in the prevalence of both PBC and AIH at more northerly latitudes; such a gradient was not apparent for PSC (Tables 2 and S3 and Figure 3). The prevalence of PBC increased by 5.61 (2.61-8.62)/100 000/degree of latitude, r^2^=0.777, p=0.003. For PSC, prevalence did not change significantly at 0.64 (*-*0.14 to 1.40)/100 000/degree of latitude, r^2^=0.406, p=0.090). For AIH, prevalence increased by 1.72 (0.36-3.08)/100 000/degree of latitude, r^2^=0.616, p=0.021.

**Figure 3:**
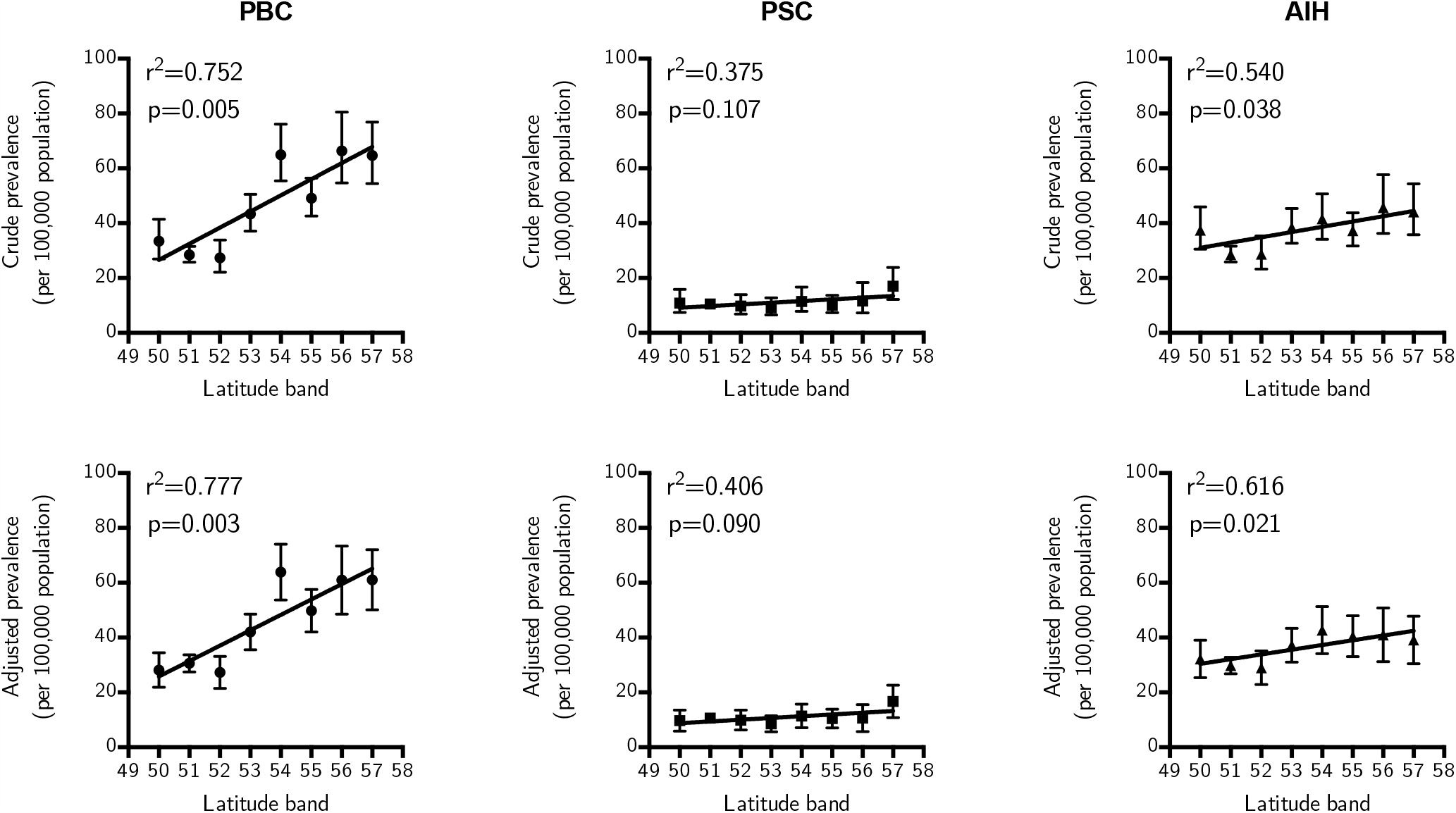
Prevalence and latitude. Crude (top row) and adjusted prevalence (bottom row) at the end of 2015 for PBC, PSC and AIH; For PBC, 5.89(2.51-9.27)/100 000/degree, r^2^=0.752, p=0.005; PSC, 0.62(−0.18-1.41)/100 000/degree, r^2^=0.375, p=0.107; AIH, 1.92(0.150-3.69)/100 000/degree, r^2^=0.540, p=0.038

Given apparent differences in incidence between latitude bands, patient sex and age at presentation were assessed by latitude band. There was a significant trend towards the younger incidence of AIH at more northerly latitudes (*-*0.87(*-*1.49 to *-*0.25) years/degree, r^2^=0.663, p=0.014), but no significant difference in age for incident PBC or PSC, and no significant difference in incident sex ratios in any disease (Tables S6 & S7).

### Sex and age

Over the whole study period, 1141 (86.8%) incident cases of PBC were female giving a female:male ratio of 6.6:1; for PSC, 230 (58.1%) were male giving a 1.4:1 male:female ratio; for AIH, 793 (76.6%) were female giving a female:male ratio of 3.3:1. There was no significant change in the sex ratio of incident cases over time for any of the conditions (Table S8). The median age of incident PBC was 63 years (IQR 53-72); for PSC 57 years (IQR 43-69 years); and for AIH 58 years (IQR 44-69). The age of incidence did not change for PBC and PSC over the study period but for AIH, median age at incidence increased (Table S9).

### Smoking

After adjustment for sex, age, Townsend deprivation quintile, and latitude, incident PBC was more frequent in smokers than those who had never smoked at 3.40 (3.03-3.77)/100 000/year and 1.96 (1.80-2.12) cases/100 000/year respectively. After the same adjustments, there was a lower incidence of PSC in smokers 0.47 (0.33-0.61)/100 000/year compared to those who had never smoked 0.95 (0.83-1.07)/100 000/year. For AIH, there was no difference between current smokers and those who had never smoked.

### Deprivation

PBC was significantly associated with deprivation. Incidence was 2.21 (1.91-2.51)/100 000/year and in the most deprived quintile 2.82 (2.35-3.28)/100 000/year after adjustment for age, sex, latitude, and smoking status. When assessed by linear regression, there was an increase in incidence of 0.14 (0.08-0.20)/100 000/year per Townsend quintile; r^2^=0.946; p=0.005. The incidence of PSC and AIH did not vary significantly with deprivation.

### Ethnicity

Data on ethnicity were missing for 5 008 804 (58.3%) registrations and a similar proportion of those diagnosed with AILD (Tables S2 & S3). Among the minority with a recorded ethnicity, the crude incidence and prevalence of PBC amongst those of white ethnicity was greater than those of non-white ethnicities. For PSC and AIH, there were no significant differences. Attempts to adjust for co-variables were not made given the high frequency of missing data.

## Discussion

In this study we use aggregate data from a nationally representative cross-section of primary care practices to show for the first time markedly greater incidences of both PBC and AIH at more northerly latitudes. For PBC there was a greater than doubling of incidence over 7 degrees of latitude; for AIH, there was an increase of over 50%. These observations persisted after correction for sex, age, deprivation, and smoking status. For PBC, the difference was notable for being more marked than the fold change in incidence for MS in either this study or in meta-analyses.[24, 25] This observation may inform investigation of aetiopathogenesis in PBC and AIH and may explain some missing disease risk and variation between others’ studies.

One potential explanation for a disease correlation with latitude is varying sunlight (ultraviolet light) exposure and its effects on vitamin D metabolism. Such a pathway has been proposed in PBC previously.[21, 34] However, if this were a major aetiological factor, those with more pigmented skin might be expected to be at elevated risk.[35] Perhaps differences in sunlight exposure modulate genetic risk. Additionally, it is unclear as to what stage in life any exposure effect from differing latitudes may have its effect and it is plausible that it is childhood exposure that is most important.[26] Expanding this work to other geographical areas is necessary. Equally, looking for similar correlations in disease incidence and latitude in other diseases associated with low vitamin D status such as inflammatory bowel disease would be valuable.[36]

Our estimates for incidence are close to those reported elsewhere: in a recent meta-analysis, the population incidence of PSC across a number of countries was estimated at 0-1.3/100 000/year and 0.33-5.8/100 000/year for PBC.[5] By comparison, in this study, overall incidence is calculated at 0.74/100 000/year and 2.47/100 000/year respectively. For AIH, meta-analyses are lacking but two recent studies from Europe report incidences of 1.1/100 000/year and 1.7/100 000/year; we report an incidence of 1.94/100 000/year.[6, 7] Our overall prevalence estimates are also similar to those published elsewhere.

There are likely to be differences in this cohort compared to others. We report median ages for the diagnosis of AILD that are higher than those reported elsewhere. For example, the median age of diagnosis of PBC in our cohort is 63 years; in the UK-PBC national cohort the median age was 55 years.[37] For PSC, our median age is 57 years, the International PSC Study Group reported a mean age at diagnosis of 39 years.[38] Such differences may reflect age-associated differences in referral patterns to, or retention of follow-up in, secondary care; bias in entry into registries; or differences in diagnostic classification between primary and secondary care. Our work has shown an apparent narrowing in the female:male ratio of patients referred for transplantation in PBC.[4] In this study, there was no change over time in the sex ratio for any disease including PBC. This may reflect the reported poorer outcome for men than women diagnosed with PBC, an observation that has in turn been related to later diagnosis.[37]

In this study we confirm the previously identified dichotomous effect of smoking on AILD risk by demonstrating an association with increased incidence of both PBC & AIH, but with decreased PSC risk. Notably, these associations persisted after controlling for deprivation, which may be associated with smoking behaviour.[39] We note negative national trends in smoking over our study period, although this study is not powered to demonstrate differences in duration of exposure and time from exposure that might be expected to affect future AILD epidemiology as national smoking trends change.[40]

Ethnicity, with its potential associations with both genetics and environment remains an incompletely understood component of AILD risk. This study was hampered by a large proportion of individuals not having a recorded ethnicity. However, we do describe an approximately doubled incidence of PBC in those of white ethnicity as compared to those of other ethnicities. Such an association has been described elsewhere, but this large cohort underlines such differences.[41] We are unable to comment on differences in likelihood of investigation for PBC in these populations nor on disease trajectory or severity at diagnosis described by others.[42]

This study has several strengths: its 14-year time frame, large cohort size, the presence on information about co-factors, and its derivation from primary care records rather than secondary care records avoids the latters’ inherent risk of selection bias. Potential weaknesses of this study include the possibility of inaccurate recording of diagnoses. We note however work suggesting that at least for AIH, primary care Read codes are broadly representative of specialist diagnoses.[31] In addition, we may have under-estimated the incidence of these diseases because of our exclusion of overlapping disease and incident disease within one year of joining a general practice. We cannot exclude the existence of an alternative confounding factor in relation to genetics, the environment, or medical practice that would explain the variations in PBC and AIH seen at different latitudes. We note specifically the existence of a centre with a particular focus on AILDs in Newcastle in the 54° latitude band. Our findings require confirmation in a different geographic region. A high proportion of individuals did not have ethnicity recorded meaning that this was excluded from analysis and ethnicity represents a further potential confounder.

Here we demonstrate for the first time a striking correlation between increased geographic latitude and disease risk for PBC; the same phenomenon is present to a lesser extent for AIH but absent for PSC. We also present key demographic information with regards to sex, age, deprivation quintile and smoking status derived from primary care data. Our results support a new avenue of investigation in AILD aetiology, provide primary-care-derived estimates of AILD epidemiology, and confirm others’ findings regarding the environmental impact of smoking, deprivation, and ethnicity on the AILDs.

## Data Availability

Data presented may be made available to other researchers in liaison with The Health Improvement Network.

## Notes

**Disclaimer and support** GJW and GMH have received support from the National Institute for Health Research Birmingham Biomedical Research Unit. GJW has benefitted from a UK Medical Research Council Clinical Research Fellowship. GMH receives further support from the UK Medical Research Council funded UK-PBC stratified medicine platform (www.uk-pbc.com), an EU Career Development Award and is also Chief Investigator for UK-PSC (www.uk-psc.com), a NIHR Translational Research Collaboration. TM is supported by the National Institute for Health Research (NIHR) Applied Research Collaboration (ARC) West Midlands. The views expressed are those of the author(s) and not necessarily those of the NIHR or the Department of Health and Social Care. This report presents independent research partly funded by the National Institute for Health Research (NIHR). The views expressed are those of the authors and not necessarily those of the NHS, the NIHR or the Department of Health. This report was partly supported by an educational grant from Intercept Pharmaceuticals.

### Competing Interest Statement

The authors have declared no competing interest.

### Funding Statement

GJW and GMH have received support from the National Institute for Health Research Birmingham Biomedical Research Unit. GJW has benefitted from a UK Medical Research Council Clinical Research Fellowship. GMH receives fur- ther support from the UK Medical Research Council funded UK-PBC stratified medicine platform (www.uk-pbc.com), an EU Career Development Award and is also Chief Investigator for UK-PSC (www.uk-psc.com), a NIHR Translational Research Collaboration. TM is supported by the National Institute for Health Research (NIHR) Applied Research Collaboration (ARC) West Midlands. The views expressed are those of the author(s) and not necessarily those of the NIHR or the Department of Health and Social Care.
This report presents independent research partly funded by the National Institute for Health Research (NIHR). The views expressed are those of the au- thors and not necessarily those of the NHS, the NIHR or the Department of Health.
This report was partly supported by an educational grant from Intercept Pharmaceuticals.

## References

[1] Gideon M Hirschfield et al. “Primary sclerosing cholangitis”. In: Lancet 382.9904 (Nov. 2013), pp. 1587–99. doi: 10.1016/S0140-6736(13)60096-3.

[2] Gideon M Hirschfield and M Eric Gershwin. “The immunobiology and pathophysiology of primary biliary cirrhosis”. In: Annu Rev Pathol 8 (Jan. 2013), pp. 303–30. doi: 10.1146/annurev-pathol-020712-164014.

[3] G J Webb et al. “Cellular and Molecular Mechanisms of Autoimmune Hepatitis”. In: Annu Rev Pathol 13 (Jan. 2018), pp. 247–292. doi: 10.1146/annurev-pathol-020117-043534.

[4] Gwilym James Webb et al. “Twenty-Year Comparative Analysis of Patients With Autoimmune Liver Diseases on Transplant Waitlists”. In: Clin Gastroenterol Hepatol 16.2 (Feb. 2018), 278–287.e7. doi: 10.1016/j.cgh.2017.09.062.

[5] Kirsten Boonstra, Ulrich Beuers, and Cyriel Y Ponsioen. “Epidemiology of primary sclerosing cholangitis and primary biliary cirrhosis: a systematic review”. In: J Hepatol 56.5 (May 2012), pp. 1181–8. doi: 10.1016/j.jhep.2011.10.025.

[6] Nicole M F van Gerven et al. “Epidemiology and clinical characteristics of autoimmune hepatitis in the Netherlands”. In: Scand J Gastroenterol 49.10 (Oct. 2014), pp. 1245–54. doi: 10.3109/00365521.2014.946083.

[7] Lisbet Gronbaek, Hendrik Vilstrup, and Peter Jepsen. “Autoimmune hepatitis in Denmark: incidence, prevalence, prognosis, and causes of death. A nationwide registry-based cohort study”. In: J Hepatol 60.3 (Mar. 2014), pp. 612–7. doi: 10.1016/j.jhep.2013.10.020.

[8] Mehul Lamba, Jing Hieng Ngu, and Catherine A M Stedman. “Trends in Incidence of Autoimmune Liver Diseases and Increasing Incidence of Autoimmune Hepatitis”. In: Clin Gastroenterol Hepatol (June 2020). doi: 10.1016/j.cgh.2020.05.061.

[9] G J Webb and G M Hirschfield. “Using GWAS to identify genetic predisposition in hepatic autoimmunity”. In: J Autoimmun 66 (Jan. 2016), pp. 25–39. doi: 10.1016/j.jaut.2015.08.016.

[10] M I Prince, S J Ducker, and O F W James. “Case-control studies of risk factors for primary biliary cirrhosis in two United Kingdom populations”. In: Gut 59.4 (Apr. 2010), pp. 508–12. doi: 10.1136/gut.2009.184218.

[11] M Eric Gershwin et al. “Risk factors and comorbidities in primary biliary cirrhosis: a controlled interview-based study of 1032 patients”. In: Hepatology 42.5 (Nov. 2005), pp. 1194–202. doi: 10.1002/hep.20907.

[12] Christophe Corpechot et al. “Demographic, lifestyle, medical and familial factors associated with primary biliary cirrhosis”. In: J Hepatol 53.1 (July 2010), pp. 162–9. doi: 10.1016/j.jhep.2010.02.019.

[13] D Howel et al. “An exploratory population-based case-control study of primary biliary cirrhosis”. In: Hepatology 31.5 (May 2000), pp. 1055–60. doi: 10.1053/he.2000.7050.

[14] Richard J Q McNally et al. “No rise in incidence but geographical heterogeneity in the occurrence of primary biliary cirrhosis in North East England”. In: Am J Epidemiol 179.4 (Feb. 2014), pp. 492–8. doi: 10.1093/aje/kwt308.

[15] K J van Erpecum et al. “Risk of primary sclerosing cholangitis is associated with nonsmoking behavior”. In: Gastroenterology 110.5 (May 1996), pp. 1503–6. doi: 10.1053/gast.1996.v110.pm8613056.

[16] S A Mitchell et al. “Cigarette smoking, appendectomy, and tonsillectomy as risk factors for the development of primary sclerosing cholangitis: a case control study”. In: Gut 51.4 (Oct. 2002), pp. 567–73. doi: 10.1136/gut.51.4.567.

[17] Sally Appleyard, Ruma Saraswati, and David A Gorard. “Autoimmune hepatitis triggered by nitrofurantoin: a case series”. In: J Med Case Rep 4 (Sept. 2010), p. 311. doi: 10.1186/1752-1947-4-311.

[18] A Gough et al. “Minocycline induced autoimmune hepatitis and systemic lupus erythematosus-like syndrome”. In: BMJ 312.7024 (Jan. 1996), pp. 169–72. doi: 10.1136/bmj.312.7024.169.

[19] Wendy Dankers et al. “Vitamin D in Autoimmunity: Molecular Mechanisms and Therapeutic Potential”. In: Front Immunol 7 (2016), p. 697. doi: 10.3389/fimmu.2016.00697.

[20] Michael F Holick. “Sunlight and vitamin D for bone health and prevention of autoimmune diseases, cancers, and cardiovascular disease”. In: Am J Clin Nutr 80.6 Suppl (Dec. 2004), 1678S–88S. doi: 10.1093/ajcn/80.6.1678S.

[21] Brian D Juran and Konstantinos N Lazaridis. “Environmental factors in primary biliary cirrhosis”. In: Semin Liver Dis 34.3 (Aug. 2014), pp. 265–72. doi: 10.1055/s-0034-1383726.

[22] Yoav Arnson, Howard Amital, and Yehuda Shoenfeld. “Vitamin D and autoimmunity: new aetiological and therapeutic considerations”. In: Ann Rheum Dis 66.9 (Sept. 2007), pp. 1137–42. doi: 10.1136/ard.2007.069831.

[23] Robyn M Lucas et al. “Vitamin D status: multifactorial contribution of environment, genes and other factors in healthy Australian adults across a latitude gradient”. In: J Steroid Biochem Mol Biol 136 (July 2013), pp. 300–8. doi: 10.1016/j.jsbmb.2013.01.011.

[24] Steve Simpson Jr et al. “Latitude is significantly associated with the prevalence of multiple sclerosis: a meta-analysis”. In: J Neurol Neurosurg Psychiatry 82.10 (Oct. 2011), pp. 1132–41. doi: 10.1136/jnnp.2011.240432.

[25] Steve Simpson Jr et al. “Latitude continues to be significantly associated with the prevalence of multiple sclerosis: an updated meta-analysis”. In: J Neurol Neurosurg Psychiatry 90.11 (Nov. 2019), pp. 1193–1200. doi: 10.1136/jnnp-2018-320189.

[26] E Hyppönen et al. “Intake of vitamin D and risk of type 1 diabetes: a birth-cohort study”. In: Lancet 358.9292 (Nov. 2001), pp. 1500–3. doi: 10.1016/S0140-6736(01)06580-1.

[27] Yinon Shapira, Nancy Agmon-Levin, and Yehuda Shoenfeld. “Defining and analyzing geoepidemiology and human autoimmunity”. In: J Autoimmun 34.3 (May 2010), J168–77. doi: 10.1016/j.jaut.2009.11.018.

[28] Betina T Blak et al. “Generalisability of The Health Improvement Network (THIN) database: demographics, chronic disease prevalence and mortality rates”. In: Inform Prim Care 19.4 (2011), pp. 251–5. doi: 10.14236/jhi.v19i4.820.

[29] Philip N Okafor et al. “Secondary analysis of large databases for hepatology research”. In: J Hepatol 64.4 (Apr. 2016), pp. 946–56. doi: 10.1016/j.jhep.2015.12.019.

[30] 2019. url: https://www.the-health-improvement-network.com/.

[31] Fumi Varyani et al. “The communication of a secondary care diagnosis of autoimmune hepatitis to primary care practitioners: a population-based study”. In: BMC Health Serv Res 13 (May 2013), p. 161. doi: 10.1186/1472-6963-13-161.

[32] Andrew Maguire, Betina T Blak, and Mary Thompson. “The importance of defining periods of complete mortality reporting for research using automated data from primary care”. In: Pharmacoepidemiol Drug Saf 18.1 (Jan. 2009), pp. 76–83. doi: 10.1002/pds.1688.

[33] Ronac Mamtani et al. “Distinguishing incident and prevalent diabetes in an electronic medical records database”. In: Pharmacoepidemiol Drug Saf 23.2 (Feb. 2014), pp. 111–8. doi: 10.1002/pds.3557.

[34] Arndt Vogel, Christian P Strassburg, and Michael P Manns. “Genetic association of vitamin D receptor polymorphisms with primary biliary cirrhosis and autoimmune hepatitis”. In: Hepatology 35.1 (Jan. 2002), pp. 126–31. doi: 10.1053/jhep.2002.30084.

[35] T Hagenau et al. “Global vitamin D levels in relation to age, gender, skin pigmentation and latitude: an ecologic meta-regression analysis”. In: Osteoporos Int 20.1 (Jan. 2009), pp. 133–40. doi: 10.1007/s00198-008-0626-y.

[36] Rita Del Pinto et al. “Association Between Inflammatory Bowel Disease and Vitamin D Deficiency: A Systematic Review and Meta-analysis”. In: Inflamm Bowel Dis 21.11 (Nov. 2015), pp. 2708–17. doi: 10.1097/MIB.0000000000000546.

[37] Marco Carbone et al. “Sex and age are determinants of the clinical phenotype of primary biliary cirrhosis and response to ursodeoxycholic acid”. In: Gastroenterology 144.3 (Mar. 2013), 560–569.e7, quiz e13–4. doi: 10.1053/j.gastro.2012.12.005.

[38] Tobias J Weismüller et al. “Patient Age, Sex, and Inflammatory Bowel Disease Phenotype Associate With Course of Primary Sclerosing Cholangitis”. In: Gastroenterology 152.8 (June 2017), 1975–1984.e8. doi: 10.1053/j.gastro.2017.02.038.

[39] Louise Marston et al. “Smoker, ex-smoker or non-smoker? The validity of routinely recorded smoking status in UK primary care: a cross-sectional study”. In: BMJ Open 4.4 (Apr. 2014), e004958. doi: 10.1136/bmjopen-2014-004958.

[40] Colin R Simpson, Julia Hippisley-Cox, and Aziz Sheikh. “Trends in the epidemiology of smoking recorded in UK general practice”. In: Br J Gen Pract 60.572 (Mar. 2010), e121–7. doi: 10.3399/bjgp10X483544.

[41] Daniel Smyk et al. “Primary biliary cirrhosis: family stories”. In: Autoimmune Dis 2011 (2011), p. 189585. doi: 10.4061/2011/189585.

[42] Marion G Peters et al. “Differences between Caucasian, African American, and Hispanic patients with primary biliary cirrhosis in the United States”. In: Hepatology 46.3 (Sept. 2007), pp. 769–75. doi: 10.1002/hep.21759.

